# Automated detection of speech timing alterations in autopsy-confirmed non-fluent/agrammatic variant primary progressive aphasia

**DOI:** 10.1101/2022.02.21.22271228

**Authors:** Adolfo M. García, Ariane E. Welch, Maria Luisa Mandelli, Maya L. Henry, Sladjana Lukic, María José Torres-Prioris, Jessica Deleon, Buddhika M. Ratnasiri, Diego L. Lorca Puls, Bruce L. Miller, William W. Seeley, Adam P. Vogel, Maria Luisa Gorno-Tempini

**Affiliations:** Global Brain Health Institute, University of California, San Francisco, CA, USA; Cognitive Neuroscience Center, Universidad de San Andrés, Buenos Aires, Argentina; National Scientific and Technical Research Council (CONICET), Buenos Aires, Argentina; Departamento de Lingüística y Literatura, Facultad de Humanidades, Universidad de Santiago de Chile, Santiago, Chile; Memory and Aging Center, Department of Neurology, University of California, San Francisco, CA, USA; Department of Communication Sciences and Disorders, University of Texas at Austin, USA; Department of Communication Sciences and Disorders, Adelphi University, Garden City, NY, USA; Cognitive Neurology and Aphasia Unit, Centro de Investigaciones Médico-Sanitarias, University of Malaga, Malaga, Spain; Instituto de Investigación Biomédica de Málaga - IBIMA, Malaga, Spain; Area of Psychobiology, Faculty of Psychology and Speech Therapy, University of Malaga, Malaga, Spain; Department of Speech, Language and Hearing Sciences, Faculty of Medicine, Universidad de Concepción, Concepción, Chile; Centre for Neuroscience of Speech, Department of Audiology & Speech Pathology, The University of Melbourne, Melbourne, Australia; Redenlab, Melbourne, Australia

**Keywords:** Non-fluent/agrammatic variant primary progressive aphasia, corticobasal degeneration, progressive supranuclear palsy, automated speech analysis, speech timing, cortical thickness, whiter matter atrophy

## Abstract

**Background and Objectives:** Motor speech function, including speech timing, is a key domain for diagnosing non-fluent/agrammatic variant primary progressive aphasia (nfvPPA). Yet, standard assessments employ subjective, specialist-dependent evaluations, undermining reliability and scalability. Moreover, few studies have examined relevant anatomo-clinical alterations in patients with pathologically-confirmed diagnoses. This study overcomes such caveats via automated speech timing analyses in a unique cohort of autopsy-proven cases.

**Methods:** In a cross-sectional study, we administered an overt reading task and quantified articulation rate, mean syllable and pause duration, and syllable and pause duration variability. Neuroanatomical disruptions were assessed via cortical thickness and white matter atrophy analysis.

**Results:** We evaluated 22 persons with nfvPPA (mean age: 67.3; 13 females) and confirmed underlying four-repeat tauopathy, 15 persons with semantic variant primary progressive aphasia (svPPA; mean age: 66.5; 8 females), and 10 healthy controls (HCs; 70 years; 5 females). All five speech timing measures revealed alterations in persons with nfvPPA relative to both the HC and svPPA groups, controlling for dementia severity. Articulation rate robustly discriminated individuals with nfvPPA from HCs (AUC = .95), outperforming specialist-dependent perceptual measures of dysarthria and apraxia of speech severity. Patients with nfvPPA exhibited structural abnormalities in left precentral and middle frontal as well as bilateral superior frontal regions, including their underlying white matter. Articulation rate was associated with atrophy of the left pars opercularis and supplementary/presupplementary motor areas. Secondary analyses showed that, controlling for dementia severity, all measures yielded greater deficits in patients with nfvPPA and corticobasal degeneration (nfvPPA-CBD, *n* = 12) than in those with progressive supranuclear palsy pathology (nfvPPA-PSP, *n* = 10). Articulation rate robustly discriminated between individuals in each subgroup (AUC = .82). More widespread cortical thinning was observed for the nfvPPA-CBD than the nfvPPA-PSP group across frontal regions.

**Discussion:** Automated speech timing analyses can capture specific markers of nfvPPA while potentially discriminating between patients with different tauopathies. Thanks to its objectivity and scalability, this approach could support standard speech assessments.

## Introduction

Non-fluent/agrammatic variant primary progressive aphasia (nfvPPA) is a clinical phenotype of frontotemporal dementia (FTD) spectrum disorders, often caused by frontotemporal lobar degeneration (FTLD). Most cases present with corticobasal degeneration (CBD) or progressive supranuclear palsy (PSP) pathology,^1-4^ two types of FTLD-four-repeat tauopathy (4Rtau).^4-7^ Both involve aberrant deposition of the microtubule-associated protein tau; varying patterns of frontal, insular, and/or striatal atrophy; underlying white matter (WM) abnormalities; and early motor speech deficits that may be accompanied by agrammatism.^4-7^ While isolated agrammatism is rare, motor speech deficits are a defining clinical feature of nfvPPA and often appear on their own –a pattern sometimes termed primary progressive apraxia of speech (PPAOS).^8^

Prominent among these deficits is abnormal speech timing, a form of dysprosody affecting the rhythm, pace, and duration of oral production. Speech timing disruptions are variant-specific markers of nfvPPA, including path-confirmed cases.^2, 9-13^ Most patients show slow articulation rate and/or prolonged syllables and extended pauses between syllables or words. Speech timing measures have thus been proposed as targets for diagnosis and follow-up.^11^ Indeed, these alterations might discriminate between CBD and PSP pathology, although the link remains unclear. Some studies report that abnormal speech timing and other prosodic disturbances are predominant in PPAOS caused by PSP,^8^ while others note their distinct presence in CBD.^2, 10^

Yet, evidence on these deficits in path-confirmed nfvPPA comes from perceptual evaluations, limiting its translational potential. Subjective impressions are prone to expertise and perceptual bias effects,^14, 15^ thereby presenting low validity and reliability.^16, 17^ Also, they are typically scored via short rating scales,^18^ bound to ceiling effects and blind to pathological differentiations. Furthermore, their administration requires trained experts, who may be unavailable across centers, especially in low-income countries.^19^

These limitations can be overcome with automated speech analysis, an objective, scalable approach which detects patterns that escape human raters^20^ and outperforms perceptual evaluations.^20^ In studies on clinically diagnosed nfvPPA, automated speech timing measures (e.g., articulation rate as well as sound and pause duration, rate, and variability) differentiate patients from healthy controls (HCs) and other PPA groups,^12, 15, 21^ capture disease progression,^21^ and correlate with phosphorylated tau level^12^ and atrophy of left motor and inferior frontal cortices.^12, 15, 21^ In particular, articulation rate is positively associated with cortical thickness of the left primary motor cortex (PMC) and supplementary/presupplementary motor areas (SMA, preSMA).^21^ Validating these measures in autopsy-proven nfvPPA would be critical to expand current assessments and discover differential markers of CBD and PSP pathology.

This study aimed to capture speech timing signatures in a well-characterized, autopsy-proven FTLD-4Rtau nfvPPA cohort relative to HCs and patients with svPPA. We administered an overt reading task;^18^ analyzed five relevant measures; assessed their correlations with specialist-dependent motor speech ratings; and examined underlying patterns of cortical thickness and WM atrophy. We hypothesized that automated speech timing measures would (a) differentiate persons with nfvPPA from both HCs and persons with svPPA, (b) robustly classify individuals with nfvPPA, and (c) reveal distinctions that are only partly captured by perceptual assessments. Moreover, we expected such deficits to correlate with atrophy of the left PMC, SMA, preSMA, and/or inferior frontal regions.^21, 22^ As a secondary aim, we explored whether speech timing deficits can discriminate between individuals with nfvPPA-CBD and nfvPPA-PSP.

## Methods

### Participants

The study involved 47 English-speaking participants completing prospective longitudinal assessments at the Memory and Aging Center (University of California, San Francisco) between 2001 and 2018 (for demographics, see Table 1; for neurological details, see eAppendix 1, eTable1). These comprised 22 persons with nfvPPA, 15 with svPPA, and 10 HCs. The study focused on persons with clinical diagnosis of nfvPPA caused by 4Rtau, with post-mortem confirmation of CBD or PSP pathology.

**Table 1.**
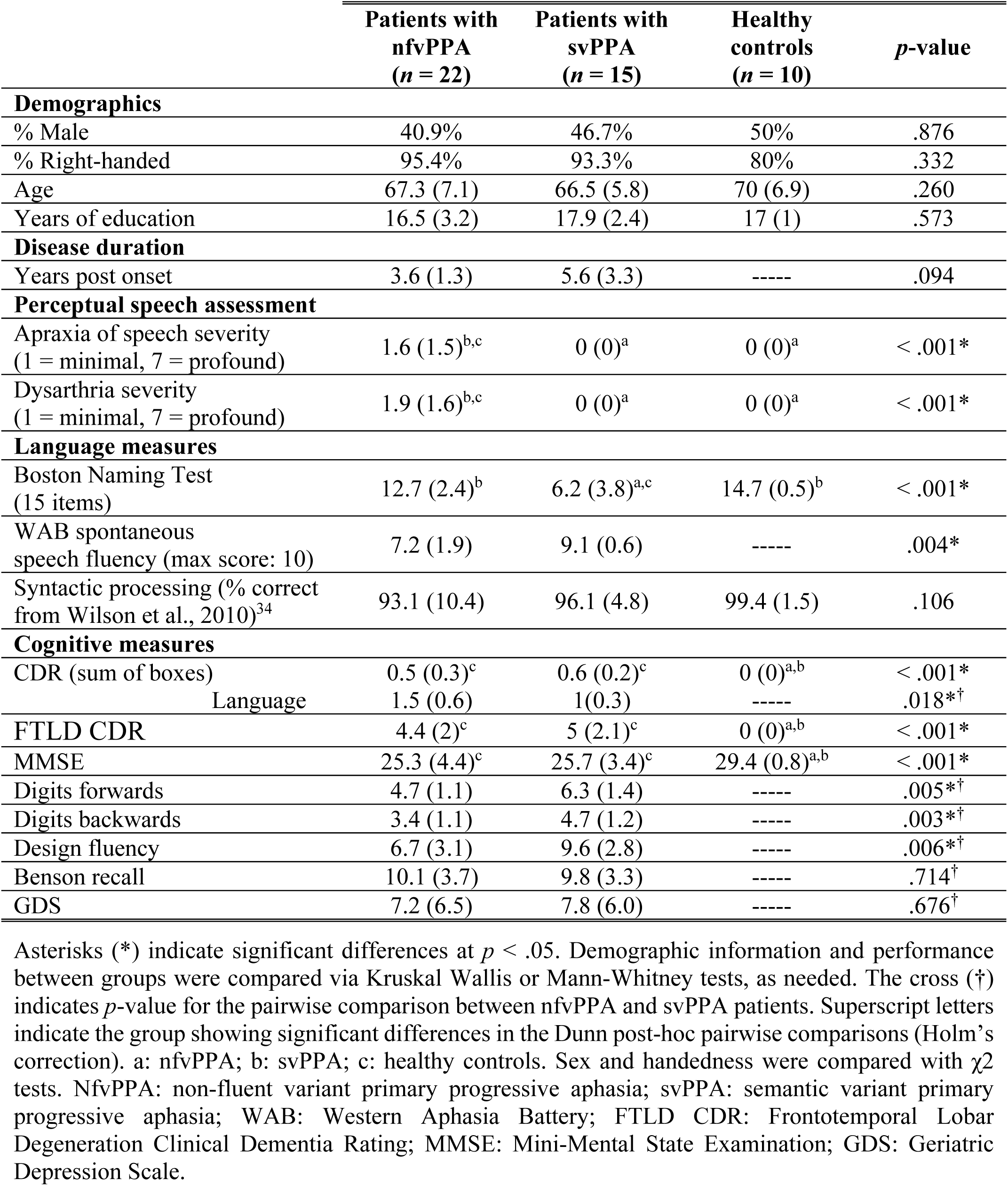
Participants’ demographic, speech, language, and cognitive profiles in the three-group setting.

|  | **Patients with**<br>**nfvPPA**<br>**(*n* = 22)** | **Patients with**<br>**svPPA**<br>**(*n* = 15)** | **Healthy**<br>**controls**<br>**(*n* = 10)** | ***p*-value** |
| --- | --- | --- | --- | --- |
| **Demographics** |  |  |  |  |
| % Male | 40.9% | 46.7% | 50% | .876 |
| % Right-handed | 95.4% | 93.3% | 80% | .332 |
| Age | 67.3 (7.1) | 66.5 (5.8) | 70 (6.9) | .260 |
| Years of education | 16.5 (3.2) | 17.9 (2.4) | 17 (1) | .573 |
| **Disease duration** |  |  |  |  |
| Years post onset | 3.6 (1.3) | 5.6 (3.3) | ----- | .094 |
| **Perceptual speech assessment** |  |  |  |  |
| Apraxia of speech severity<br>(1 = minimal, 7 = profound) | 1.6 (1.5)<sup>b,c</sup> | 0 (0)<sup>a</sup> | 0 (0)<sup>a</sup> | < .001* |
| Dysarthria severity<br>(1 = minimal, 7 = profound) | 1.9 (1.6)<sup>b,c</sup> | 0 (0)<sup>a</sup> | 0 (0)<sup>a</sup> | < .001* |
| **Language measures** |  |  |  |  |
| Boston Naming Test<br>(15 items) | 12.7 (2.4)<sup>b</sup> | 6.2 (3.8)<sup>a,c</sup> | 14.7 (0.5)<sup>b</sup> | < .001* |
| WAB spontaneous<br>speech fluency (max score: 10) | 7.2 (1.9) | 9.1 (0.6) | ----- | .004* |
| Syntactic processing (% correct<br>from Wilson et al., 2010)<sup>34</sup> | 93.1 (10.4) | 96.1 (4.8) | 99.4 (1.5) | .106 |
| **Cognitive measures** |  |  |  |  |
| CDR (sum of boxes) | 0.5 (0.3)<sup>c</sup> | 0.6 (0.2)<sup>c</sup> | 0 (0)<sup>a,b</sup> | < .001* |
| Language | 1.5 (0.6) | 1(0.3) | ----- | .018*<sup>†</sup> |
| FTLD CDR | 4.4 (2)<sup>c</sup> | 5 (2.1)<sup>c</sup> | 0 (0)<sup>a,b</sup> | < .001* |
| MMSE | 25.3 (4.4)<sup>c</sup> | 25.7 (3.4)<sup>c</sup> | 29.4 (0.8)<sup>a,b</sup> | < .001* |
| Digits forwards | 4.7 (1.1) | 6.3 (1.4) | ----- | .005*<sup>†</sup> |
| Digits backwards | 3.4 (1.1) | 4.7 (1.2) | ----- | .003*<sup>†</sup> |
| Design fluency | 6.7 (3.1) | 9.6 (2.8) | ----- | .006*<sup>†</sup> |
| Benson recall | 10.1 (3.7) | 9.8 (3.3) | ----- | .714<sup>†</sup> |
| GDS | 7.2 (6.5) | 7.8 (6.0) | ----- | .676<sup>†</sup> |

Asterisks (\*) indicate significant differences at $p < .05$ . Demographic information and performance between groups were compared via Kruskal Wallis or Mann-Whitney tests, as needed. The cross (†) indicates $p$ -value for the pairwise comparison between nfvPPA and svPPA patients. Superscript letters indicate the group showing significant differences in the Dunn post-hoc pairwise comparisons (Holm's correction). a: nfvPPA; b: svPPA; c: healthy controls. Sex and handedness were compared with $\chi^2$ tests. NfvPPA: non-fluent variant primary progressive aphasia; svPPA: semantic variant primary progressive aphasia; WAB: Western Aphasia Battery; FTLD CDR: Frontotemporal Lobar Degeneration Clinical Dementia Rating; MMSE: Mini-Mental State Examination; GDS: Geriatric Depression Scale.

Clinical nfvPPA diagnoses were made by a multidisciplinary team including neurologists, neuropsychologists, and speech-language pathologists, following validated criteria.^23^ In all cases, speech production challenges were the first and main complaint as well as the primary cause of functional impairment. Subtle grammatical deficits were noted in most patients. Postmortem, these individuals were autopsied at the University of California, San Francisco, the University of Pennsylvania, or the Vancouver General Hospital. Pathological diagnoses were made following consensus FTLD guidelines^24^ and standard procedures.^25^ FTLD-4R-tau analysis revealed that primary pathological diagnosis was CBD for 12 individuals and PSP for the remaining 10. Following tau immunohistochemistry, all CBD patients exhibited astrocytic plaques and threadlike inclusions,^26^ while those with PSP presented globose tangles and tufted astrocytes.^27^ These subgroups had partly distinct clinical profiles (Supplement, eAppendix 2) and they were sociodemographically matched (Supplement, eAppendix 3).

The remaining participants comprised 10 HCs (with normal neurological, cognitive, speech, and language profiles) and 15 persons with svPPA (included to establish the syndromic specificity of predicted deficits). The study’s goal did not include assessments of logopenic variant PPA, although sufficient recordings will be available for future reports. Persons with svPPA were diagnosed following the abovementioned consensus criteria. No pathological information was required for these patients to enter the study, although we note that TDP Type C pathology is most common in our historical cohort. They exhibited naming and word comprehension deficits with preserved grammar and motor speech. These two groups were matched to the overall nfvPPA sample (Table 1) and its pathologically-defined subgroups (Supplement, eAppendix 3, eTable 2) in terms of sex, age, years of education, and handedness. Our sample size reached a power of 0.93 for the three-group analyses (nfvPPA, HCs, svPPA) and 0.90 for the four-group analyses (nfvPPA-CBD, nfvPPA-PSP, HCs, svPPA) –for details, see Supplement, eAppendix 4.

Perceptual assessments of videotaped sessions by certified speech pathologists, based on the validated Motor Speech Evaluation,^18^ indicated that all patients with nfvPPA exhibited dysarthria (with mixed spastic, flaccid, and/or hypokinetic speech features)^18^ and/or apraxia of speech (with abnormal articulation characterized by slow speech rate, sound distortions, and sequencing errors).^28^ Clinical ratings of dysarthria or apraxia of speech severity did not differ significantly between the nfvPPA-CBD and nfvPPA-PSP subgroups. No signs of dysarthria or apraxia of speech were observed in HCs or patients with svPPA. Results of these assessments are presented alongside linguistic and cognitive results in Table 1 and in Supplement, eAppendix 3 (eTable 2).

### Standard Protocol Approvals, Registrations, and Patient Consents

All participants or their caregivers provided written informed consent before inclusion, in accordance with the Declaration of Helsinki. The UCSF Human Research and Protection Program Institutional Review Board approved the three studies participants consented to (10-03946, 10-00619, 12-10512).

### Automated Speech Assessment

All participants performed an overt reading task, using the Grandfather Passage (Supplement, eAppendix 5). This 129-word text was designed to elicit all phonemes and main phoneme clusters in English.^18^ Unlike spontaneous or semi-spontaneous speech tasks, overt reading tasks keep verbal information constant across participants, circumventing potential confounds related to linguistic demands. Moreover, they prove sensitive to speech timing alterations in nfvPPA and other neurodegenerative conditions.^15, 29^ Participants were asked to read at their own pace, with normal volume and cadence. Their speech was recorded, saved as .wav files (44.1 KHz, 16 bits), and analyzed to capture five timing measures: *articulation rate* (syllables per second of phonated time, upon pause removal), *syllable duration* (mean across all syllables), *pause duration* (mean across all pauses, defined as silent intervals between speech sound offset and onset), *syllable duration variability* (standard deviations of syllabic durations), and *pause duration variability* (standard deviation of pause durations).

Acoustic features were extracted using onset detection functions and custom MATLAB scripts. Onset and offset detection was based on acoustic power and summed spectral energy measures across frequencies between 20-4000 Hz, using validated methods adapted here^30^ and at The University of Melbourne. Time series of these features were smoothed within 50-ms windows and normalized to yield values between 0 and 1. Time points above or below 0.1 were considered onsets or offsets. An 80-ms threshold between offset/onsets was used to limit inclusion of intra-syllabic pauses.

### Analysis of Speech Features

Speech timing variables were compared among groups via factorial ANCOVAs, covarying for MMSE score as a measure of dementia severity –as in previous work.^31^ Widely used in PPA research,^4, 5, 9, 12, 15, 18, 22, 32-34^ the MMSE was chosen over the CDR and the FTLD CDR as these two measures have a restricted range of possible scores, little variance across our (fairly mild) patients, and scores of 0 (null variability) across HCs, thus proving suboptimal as potential covariates. Analyses were conducted in a three-group setting (nfvPPA, HCs, svPPA) and in a four-group setting (nfvPPA-CBD, nfvPPA-PSP, HCs, svPPA). For each variable, participants with values ≥ 2.5 *SDs* from the group’s mean were removed as outliers. Data were excluded from (a) a single HC for three variables (syllable duration variability, pause duration, pause duration variability) in both the primary and secondary analyses; and (b) a single patient with nfvPPA-PSP for a single variable, only in the secondary analyses. No further participants had outlier values in any analysis. Missing data represented < 5% in each group for primary analyses and it was restricted to a single person with nfvPPA. Significant effects (*p* < .05) were further analyzed via Tukey’s HSD post-hoc tests. Effect sizes were calculated with η_p_^2^ for ANCOVAs and Cohen’s *d* for pairwise comparisons.

Moreover, we ran binary logistic regressions to examine whether speech timing features could classify (a) individuals with nfvPPA from HCs and (b) persons with nfvPPA-CBD from those with nfvPPA-PSP. To overcome multicollinearity issues, the most sensitive automated measure across both groups (as seen in effect sizes) was used as a single predictor. For comparison, we also ran regression models including perceptual scores of apraxia of speech and dysarthria (Table 1) as separate predictors. Classification accuracy was evaluated considering the area under the ROC curve (AUC). For exploratory purposes, we used Pearson’s *r* to examine correlations between each automated measure and specialist-dependent perceptual ratings of apraxia of speech and dysarthria severity in the overall nfvPPA sample. Analyses were run on R (R Core Team, 2020) and JASP v.0.14.1 (JASP Team, 2020).

### Neuroimaging Analyses

#### Data Acquisition and Preprocessing

All participants underwent whole-brain structural MRI on a 1.5T Siemens, 3T Siemens Trio or 3T Siemens Prisma scanners, with a T1-weighted 3D magnetization-prepared rapid-acquisition gradient echo sequence. Standard parameters were used for the 1.5T scanner (164 coronal slices; voxel size = 1.0×1.5×1.0 mm^3^; FoV = 256×256 mm^2^; matrix size = 256×256; TR = 10 ms; TE = 4 ms; T1 = 300 ms; flip angle = 15°), the 3T Trio scanner (160 sagittal slices; voxel size = 1.0×1.0×1.0 mm^3^; FoV = 256×256 mm; matrix size = 256×256; TR = 2300 ms; TE = 2.98 ms; flip angle = 9°), and the 3T Prisma scanner (160 sagittal slices; voxel size = 1.0×1.0×1.0 mm^3^; FoV = 256×256 mm; matrix size = 256×256; TR = 2300 ms; TE = 2.90 ms; flip angle = 9°). MRI data for patient groups were compared with those of a group of HCs (matched for sex, age, education, and scanner type) selected from the Hillblom Aging Network Project.

#### Cortical Thickness and White Matter Atrophy Analysis

T1-weighted images were visually inspected to ensure the absence of artifacts or excessive motion. Images were processed through the Computational Anatomy Toolbox (http://dbm.neuro.uni-jena.de/cat) in Statistical Parametric Mapping software (http://www.fil.ion.ucl.ac.uk/spm/software/spm12) running under MATLAB. Images were bias-field corrected with a spatial adaptive non-local means denoising filter^35^ and segmented into gray matter (GM), WM, and cerebrospinal fluid (CSF). All these tissue classes underwent local intensity transformation before the final adaptive maximum a posteriori segmentation,^36^ refined by applying a partial volume.^37^ Segmented images were spatially normalized to the Montreal Neurological Institute space using geodesic shooting registrations^38^ and modulated by scaling with the amount of volume changes due to spatial registration. WM images were smoothed for a voxel-based morphometry analysis with an 8-mm full-width at half-maximum Gaussian kernel.

Cortical thickness was estimated via surface-based morphometry, a measure that surpasses standard voxel-based analysis as brain surface meshes increase brain registration accuracy.^39^ To this end, the skull-stripped brain was parcellated into left and right hemisphere, subcortical regions, and cerebellum. Cortical thickness estimation and reconstruction of the central surface were performed using a projection-based thickness method.^40^ Final maps were resampled and smoothed using a 15-mm Gaussian heat kernel.^41^ Whole-brain analyses of between-group differences in cortical thickness and WM volume were performed via ANOVAs, including sex, age, handedness, and scanner type as nuisance variables. Total intracranial volume (TIV) was included for WM voxel-based morphometry but not for cortical thickness analysis, given that head size is associated only with the former measure.^42^ Alpha levels were set at *p* < .05. Family-wise error (FWE) correction was used to detect areas of peak cortical thinning and of WM volume loss. For better visualization, of between-group comparisons were also performed with a less stringent threshold (*p* < .001, uncorrected).

#### Brain-Behavior Analyses

Considering previous resarch,^21^ we assessed whether articulation rate was associated with cortical atrophy in our target ROIs. To this end, we performed linear regressions with sex, age, and TIV as covariates. We used a liberal uncorrected threshold (*p* < .05). While this is suitable given our moderate sample size and hypothesis-driven analyses,^21^ we report adjusted R-squares values, controlling for the number of terms in the model. We targeted the following left-hemisphere ROIs in specific Brodmann areas (BAs): PMC (BA4), SMA (BA6), and preSMA (BA8). We also included additional ROIs over the inferior frontal gyrus (IFG), namely: pars opercularis (BA44) and pars triangularis (BA45). All ROIs were based on the Human Brainnetome Atlas.^43^

#### Data Availability

Public archiving of anonymized data is not contemplated by the study’s IRB approval. Specific requests can be submitted through the UCSF MAC Resource Request form. Following a UCSF-regulated procedure, access will be granted to designated individuals in line with ethical guidelines on the reuse of sensitive data. This would require submission of a Material Transfer Agreement. Commercial use will not be approved.

## Results

### Automated Speech Analysis Results

#### ANCOVA Results

In the three-group setting (collapsing all patients with nfvPPA), the five speech timing measures yielded significant effects of group, adjusting for MMSE scores. Post-hoc analyses consistently revealed impaired motor speech in the nfvPPA group relative to both HCs and patients with svPPA, there being no significant differences between the latter two groups (Figure 1 and Supplement, eAppendix 6, eTable 3).

**Figure 1.**
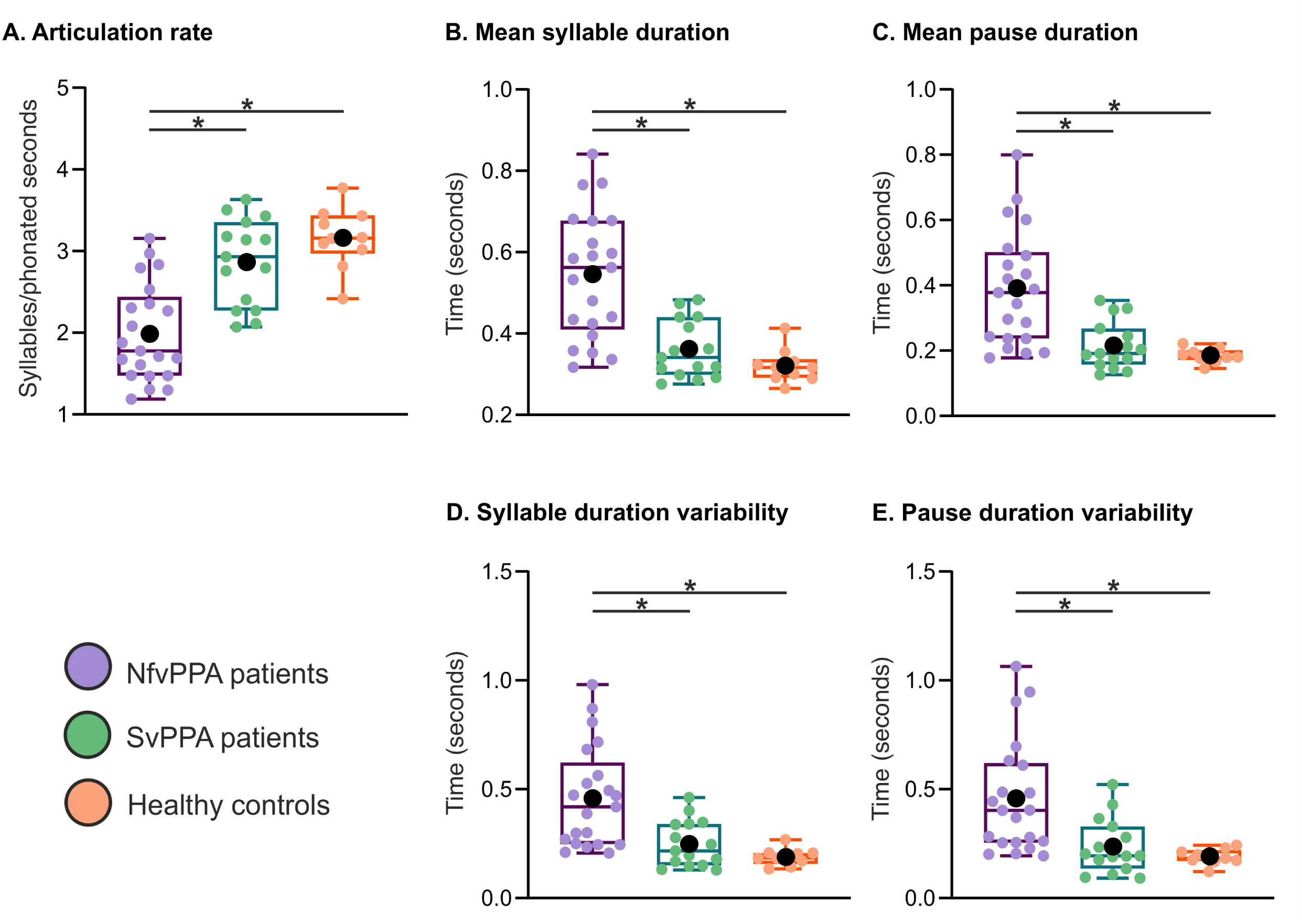
Results for the three-group setting (all nfvPPA patients together). All five speech timing measures revealed significant impairments in nfvPPA patients relative to both healthy controls and svPPA patients. No differences were observed between the latter two groups. In the box plot, middle horizontal lines show each group’s median, with lower and upper lines representing the 25^th^ and 75^th^ percentiles, respectively. The whiskers represent the smallest and largest values in the distribution. Colored dots indicate each participant’s individual value. Black dots represent the group’s mean. The asterisk (*) denotes significant differences at *p* < .05. All statistics were calculated after outlier removal (namely, a single nfvPPA-PSP participant). NfvPPA: non-fluent variant primary progressive aphasia; svPPA: semantic variant primary progressive aphasia.

Results from the four-group analysis revealed that abnormal speech timing in the nfvPPA group was primarily driven by patients with nfvPPA-CBD, who differed from HCs and patients with svPPA across all five automated measures. Conversely, deficits in the nfvPPA-PSP group were confined to articulation rate. Moreover, all five measures showed greater deficits in the nfvPPA-CBD than in the nfvPPA-PSP group, adjusting for MMSE scores (Figure 2 and Supplement, eAppendix 6, eTable 4).

**Figure 2.**
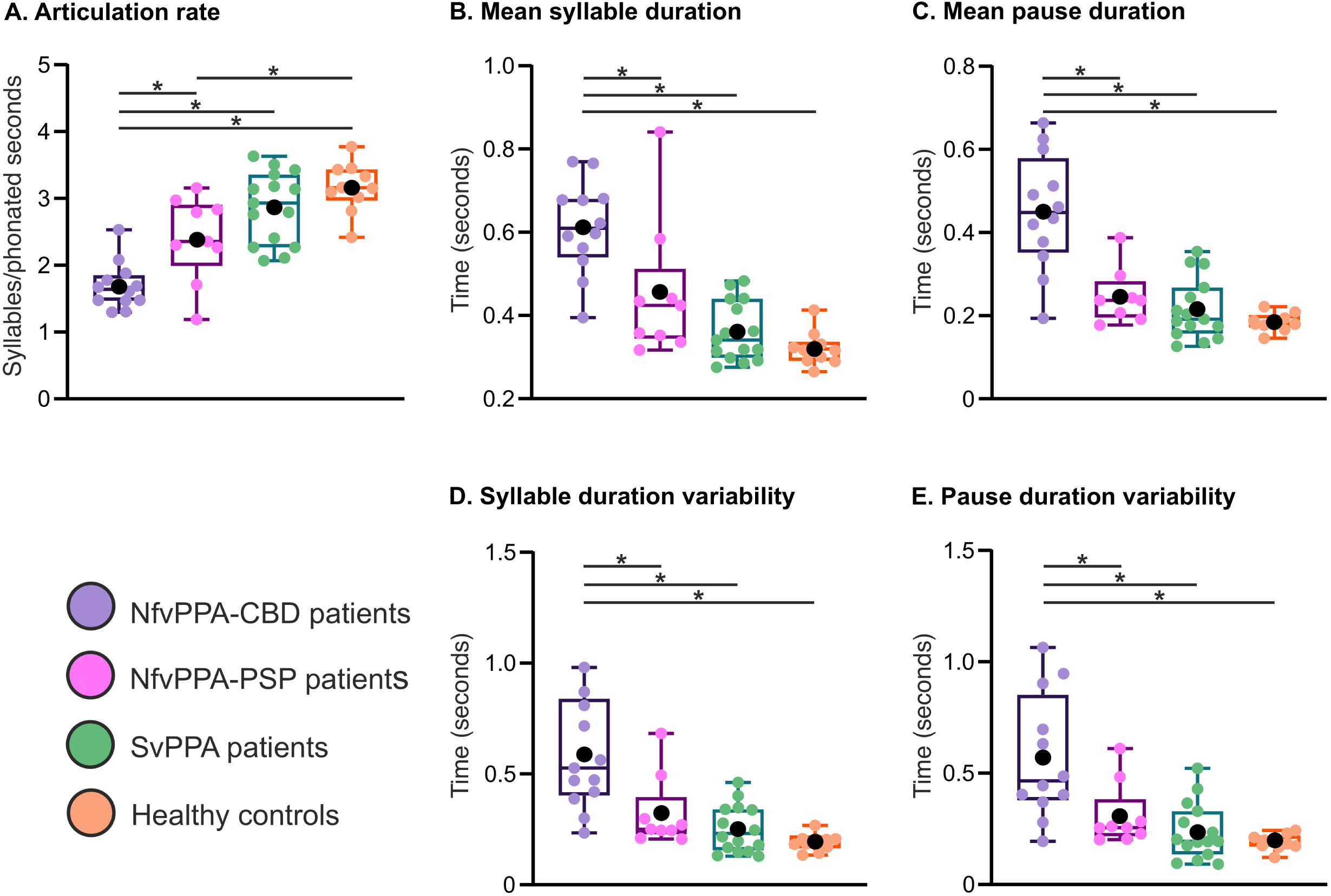
Results of the four-group analysis (separating nfvPPA-CBD from nfvPPA-PSP patients). All five speech timing measures captured significant impairments in nfvPPA-CBD patients compared to both healthy controls and svPPA patients. In contrast, only articulation rate was impaired in the nfvPPA-PSP patients. Deficits in all measures were significantly greater for nfvPPA-CBD than nfvPPA-PSP patients. In the box plot, middle horizontal lines show each group’s median, with lower and upper lines representing the 25^th^ and 75^th^ percentiles, respectively. The whiskers represent the smallest and largest values in the distribution. Colored dots indicate each participant’s individual value. Black dots represent the group’s mean. Asterisks (*) denote significant differences at *p* < .05. All statistics were calculated after outlier removal (namely, a single nfvPPA-PSP participant). NfvPPA-CBD: non-fluent variant primary progressive aphasia with corticobasal degeneration pathology; nfvPPA-PSP: non-fluent variant primary progressive aphasia with progressive supranuclear palsy pathology; svPPA: semantic variant primary progressive aphasia.

#### Binary Logistic Regression Results

Binary logistic regressions were run based on articulation rate, namely, the measure yielding the largest effect size in the three-group analyses and the only one revealing deficits in both nfvPPA subgroups. The logistic regression model showed that this predictor discriminated between patients with nfvPPA and HCs [Wald χ2 (1) = 6.53, *p* = .012] with high accuracy (AUC = .95). This single-predictor model correctly classified 90% of persons with nfvPPA and 80% of HCs. Group membership was not predicted when based on independent models of apraxia of speech [Wald χ2 (1) = 1.60, *p* = .99] or dysarthria [Wald χ2 (1) = 0, *p* = .99] severity ratings, both yielding less accurate classification than the articulation rate model (AUC = .88 and .86, respectively) –Figure 3A.

**Figure 3.**
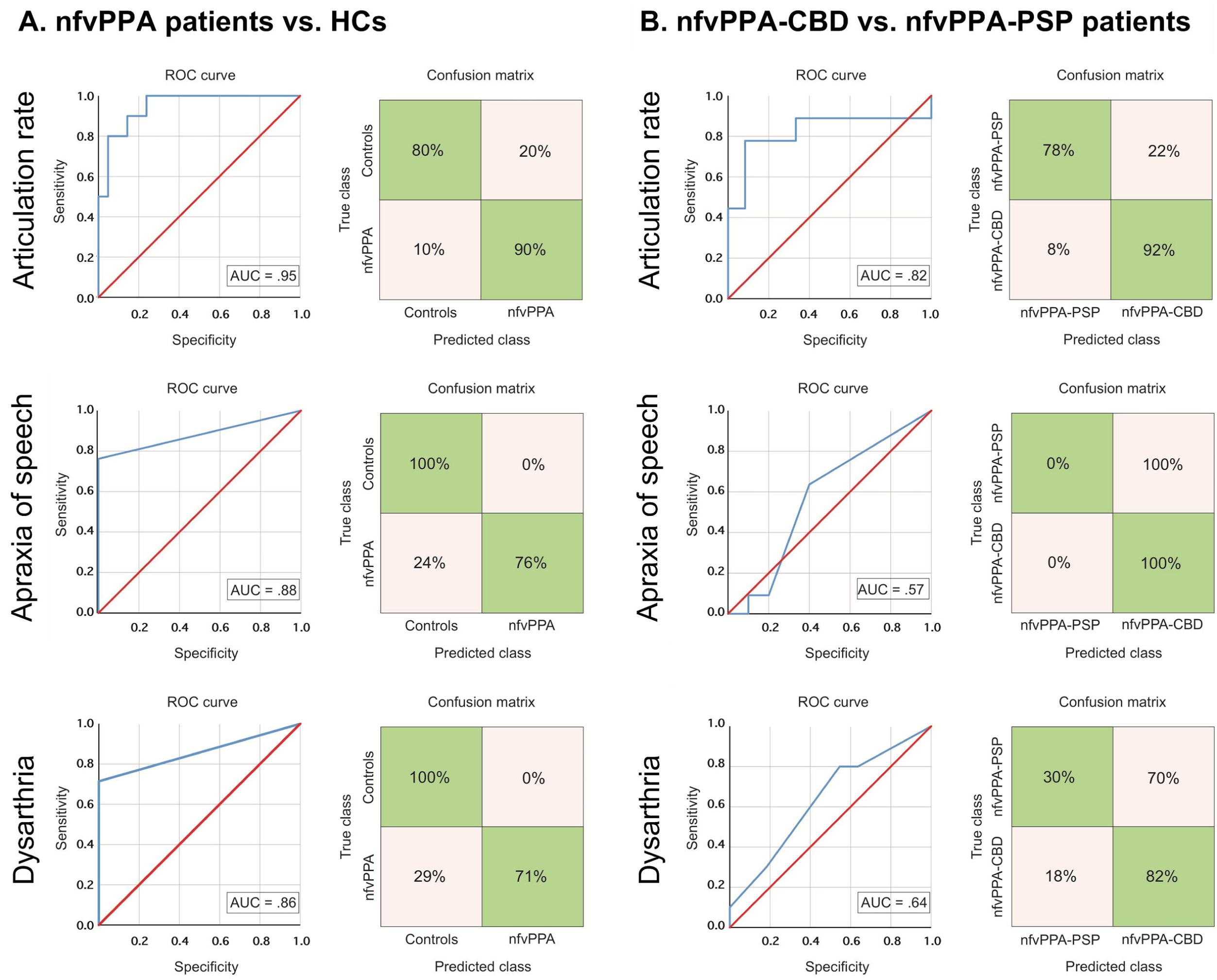
Binary logistic regression results. **(A)** Classification between nfvPPA patients and HCs was significant for the automated measure of articulation rate (top inset) as well as the perceptual measures of apraxia of speech severity (middle inset) and dysarthria severity (bottom inset), with maximal AUC score for the former. **(B)** Classification between nfvPPA-CBD and nfvPPA-PSP patients was significant for the automated measure of articulation rate (top inset) but not for the perceptual measures of apraxia of speech severity (middle inset) and dysarthria severity (bottom inset), the former yielding a robust AUC value. ROC: receiving operating characteristic; AUC: area under the ROC curve; nfvPPA: non-fluent variant primary progressive aphasia; nfvPPA-CBD: non-fluent variant primary progressive aphasia with corticobasal degeneration pathology; nfvPPA-PSP: non-fluent variant primary progressive aphasia with progressive supranuclear palsy pathology; svPPA: semantic variant primary progressive aphasia; HCs: healthy controls.

Also, articulation rate discriminated between pathologically-defined subgroups [Wald χ2 (1) = 5.51, *p* = .019]. This single-predictor model yielded high classification accuracy (AUC = .82), correctly identifying 92% of persons with nfvPPA-CBD and 78% of persons with nfvPPA-PSP. Conversely, apraxia of speech severity did not predict group membership [Wald χ2 (1) = 0.003, *p* = .954], correctly identifying 100% of persons with nfvPPA-CBD but 0% of persons with nfvPPA-PSP (AUC = .57). Likewise, group membership was not predicted by dysarthria severity [Wald χ2 (1) = 1.20, *p* = .273], identifying 82% of persons with nfvPPA-CBD and only 30% of persons with nfvPPA-PSP (AUC = .64) –Figure 3B.

### Correlations Between Automated and Perceptual Speech Measures

Across all patients with nfvPPA, perceptual ratings of apraxia of speech severity correlated negatively with articulation rate (*p* = .04, *r* = -0.45) and positively with mean syllable duration (*p* = .04, *r* = 0.46). Non-significant results were observed for every other correlation between speech timing measures and perceptual ratings of apraxia of speech and dysarthria severity (all *p*-values > .19). For details, see Supplement, eAppendix 7, eTable 5.

### Cortical Thinning and WM Loss Across Patient Groups

The combined nfvPPA group exhibited cortical thinning in the left precentral and caudal/rostral middle frontal cortices as well as the bilateral superior frontal gyrus (including the supplementary motor area), together with atrophy in the underlying WM (*p* < .05, FWE-corrected) –Figure 4A. Patients with svPPA exhibited reduced cortical thickness in the left temporal pole (extending onto inferior, middle, and superior temporal gyri) and lower volume in the underlying WM (*p* < .05, FWE-corrected) –Figure 4B. Secondary analyses in the nfvPPA-CBD and nfvPPA-PSP subgroups revealed structural abnormalities in the same areas as the combined nfvPPA group, with the nfvPPA-CBD subgroup showing more involvement and additional thinning of the left pars opercularis (*p* < .05, FWE-corrected) relative to HCs; and the nfvPPA-PSP subgroup exhibiting compromise of the angular gyrus as well as the bilateral dentate nucleus, the left thalamus, and the left midbrain (though not surviving FWE correction) relative to HCs. Structural abnormality was more widespread in patients with CBD than in those with PSP (Figure 4C-D).

**Figure 4.**
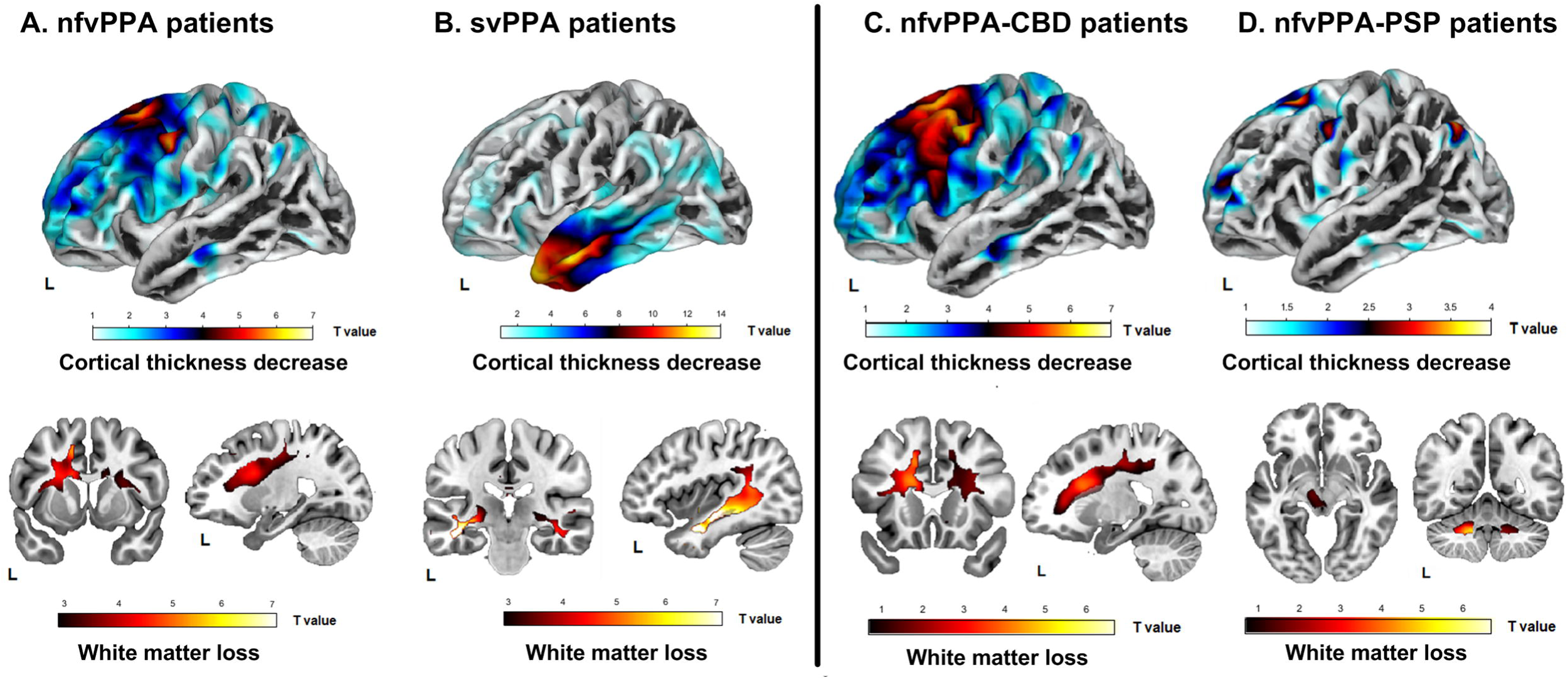
Brain structural abnormality patterns of each patient group. The combined nfvPPA group (panel **A**) and the svPPA group (panel **B**), as well as subgroups with nfvPPA-CBD (panel **C**) and nfvPPA-PSP (panel **D**), were compared with HCs to estimate their patterns of cortical thickness (top insets) and white matter volume (bottom insets) via surface-based and voxel-based morphometry, respectively. For better visualization of differences and similarities across groups, *t*-map values are reported at an uncorrected *p* < .001 threshold. NfvPPA: non-fluent variant primary progressive aphasia; nfvPPA-CBD: non-fluent variant primary progressive aphasia with corticobasal degeneration pathology; nfvPPA-PSP: non-fluent variant primary progressive aphasia with progressive supranuclear palsy pathology; svPPA: semantic variant primary progressive aphasia.

### Brain-Behavior Associations

Across the nfvPPA group, articulation rate was associated with cortical atrophy in the left SMA (*p* = .03, adjusted *R*^*2*^ = 0.18), preSMA (*p* = .02, adjusted *R*^*2*^ = 0.34), and pars opercularis (*p* < .01, adjusted *R*^*2*^ = 0.47), but not in the PMC (*p* = .07, adjusted *R*^*2*^ = 0.14) or the pars triangularis (*p* = .65, adjusted *R*^*2*^ = -0.07).

### Classification of Evidence

This study provides Class III evidence that objective speech timing measures can identify persons with autopsy-confirmed nfvPPA and discriminate between those with CBD and PSP pathology.

## Discussion

We employed automated speech timing analysis to identify patients with autopsy-confirmed nfvPPA. All five measures discriminated these individuals from HCs and from patients with svPPA, and articulation rate surpassed specialist-dependent perceptual evaluations in distinguishing individuals with nfvPPA. Abnormal speech timing was accompanied by GM and WM alterations in left frontal cortices, and significantly associated with motor region atrophy. Also, relative to patients with nfvPPA-PSP, those with nfvPPA-CBD exhibited greater deficits in all automated metrics and more widespread frontal atrophy. Finally, articulation rate robustly discriminated between patients in each subgroup. These findings suggest that automated speech timing measures are sensitive tools for nfvPPA assessments.

Relative to HCs and patients with svPPA, the nfvPPA group produced fewer syllables per phonated second as well as longer and less isochronous syllables and pauses. While variant-specific alterations of speech timing and other prosodic dimensions have been reported in clinically diagnosed nfvPPA,^11, 12^ our study successfully captured them automatically in autopsy-confirmed patients. This finding extends evidence from perceptual measures in persons with confirmed FTLD-tauopathy,^5^ indicating that speech timing markers are critically linked to such pathology.^12^ Notably, these distinctions proved significant upon controlling for dementia severity, which is notable given that cognitive impairment can influence motor speech deficits.^29^

Articulation rate (our most sensitive automated measure) classified individuals with nfvPPA from HCs with an AUC of .95. This result surpassed the highest AUC scores in the nfvPPA literature, based on both single and combined motor speech measures.^12^ As observed in other diseases,^20, 44^ the classifier based on articulation rate outperformed those based on perceptual measures. Though useful when administered by specialists, listener-based assessments may yield inconsistent results and overlook fine-grained speech dimensions.^16, 17^ The latter point may account for the better performance of our automated tools.

Both articulation rate and mean syllable duration were correlated with apraxia of speech (but not with dysarthria) severity ratings. This might reflect the prominence of phonated segment length (as opposed to pausing) in perceptual assessments, as both automated measures lean heavily on such a factor. Still, no other correlation between automated and perceptual measures reached significance, suggesting that they capture partly different phenomena. Briefly, regression results support the value of automated speech timing assessments for nfvPPA,^11^ showing that subtle dysfunctions can be objectively established at a probabilistic single-patient level.

The nfvPPA group exhibited cortical thinning across left precentral, left middle frontal, and bilateral superior frontal regions, including the SMA –alongside atrophy of the underlying WM. These regions (in particular, the precentral and supplementary motor cortices) are central hubs of the motor speech network,^32^ likely accounting for the patients’ behavioral profile. Moreover, WM alterations beneath these and other substrates are typical of nfvPPA with FTLD-tau.^33^ In fact, damage to frontal regions, especially including the SMA, is typically observed in non-agrammatic patients with motor speech disorders.^8^ Interestingly, while prosodic alterations may also involve non-cortical degeneration (e.g., in the superior cerebellar peduncle),^45^ our study suggests that at least some such disruptions (namely, speech timing alterations) may become significant without marked subcortical damage.

Articulation rate was associated with atrophy in the left SMA and preSMA, as observed by Cordella et al. (2019).^21^ However, unlike that work, we also observed an association with atrophy of the left pars opercularis, while failing to find associations with PMC or the pars triangularis. These differences may reflect the fact that brain-behavior correlations in Cordella et al. (2019)^21^ were conducted upon collapsing patients from all three PPA variants, each presenting distinct atrophy patterns.

As seen in secondary analyses, all automated measures revealed greater deficits in nfvPPA-CBD than in nfvPPA-PSP –the latter group, indeed, was impaired only on articulation rate. This measure classified persons in each subgroup with an AUC of .82 – unlike perceptual speech measures, which failed to discriminate between groups. Interestingly, a previous study found that prosodic deficits were more strongly related to PSP than CBD pathology.^8^ Yet, that study targeted (semi)spontaneous speech while we used overt reading. These tasks’ discrepant demands elicit distinct predominant motor speech disruptions, with prosodic alterations proving more salient during reading.^29^ Also, no specific assessment of speech timing was conducted therein. Furthermore, dysprosodic signs have been highlighted in CBD,^2, 10^ and slow syllabically segmented prosody is a diagnostic criterion^46^ for a speech-predominant disorder more likely caused by CBD than PSP.^47^ Thus, automated timing assessments could capture distinct signatures of nfvPPA-CBD that may be underestimated in listener-based studies.

Neuroimaging results inform these differentiations. Structural abnormality along premotor speech regions and underlying WM was present in both subgroups, as previously observed.^8, 45^ Yet, such patterns proved markedly more pronounced and widespread in persons with nfvPPA-CBD, potentially accounting for their more severe speech timing impairments. This finding opens new avenus to investigate the role of neocortical motor-network hubs in speech timing.

This study underscores the usefulness of automated acoustic measures for nfvPPA assessments. Perceptual evaluations may not always be valid or reliable,^14-17^ they are not well suited to monitoring change,^48^ and their optimal administration may be unfeasible in low-income countries lacking specialized staff.^19^ Conversely, computerized systems offer an affordable and objective framework that can be applied remotely. In particular, unlike dimensions like loudness or pitch, speech timing measures are robust to variability in noise and recording conditions, highlighting their scalability. By corroborating their sensitivity in autopsy-confirmed patients, this study addresses current calls to complement standard protocols with cutting-edge metrics.

Moreover, the potential to discriminate between CBD and PSP pathology holds clinical promise. Each of those tauopathies may be prodromal to either corticobasal syndrome (typified by asymmetric movement abnormalities, myoclonus, and dystonia) or PSP syndrome (with symmetric motor symptoms and vertical supranuclear palsy),^2, 10^ whose differentiation in early clinical testing is very challenging. Also, CBD and PSP pathology might entail distinct tau prion conformations,^49^ potentially requiring different therapies.^5^ Thus, these and other tools enabling automatic identification of nfvPPA-CBD and nfvPPA-PSP may optimize individualized treatment, prognosis, and monitoring. Note, however, that the reported measures may only discriminate between these subgroups if administered in early stages, as both will likely become more pervasive, less distinguishable speech deficits as disease progresses.

Finally, some researchers do not consider that isolated motor speech deficits constitute aphasia, and they use the term PPAOS instead. Still, current criteria indicate that clinical nfvPPA diagnosis is warranted even if patients exhibit core motor speech deficits without agrammatism and complex sentence comprehension deficits. Moreover, not all patients with predominant motor speech deficits are characterized by apraxia of speech (as the PPAOS label would suggest), but rather by dysarthria or some combination therefrom. Also, subtle grammatical difficulties were observed in our cohort during clinical interviews, supporting the view that nfvPPA is a spectrum, as the weight of motor speech and grammatical deficits varies across patients and throughout time. Future studies should address these definitional questions in greater depth.

### Limitations and avenues for further research

Our study is not without limitations. First, although we assembled the largest autopsy-confirmed cohort in the automated speech analysis literature, our sample size was modest. Future replications should involve more participants, especially for subgroup analyses. Second, inconsistent recording conditions (e.g., divergent volume and noise levels) over our 17-year data collection period precluded analysis of other acoustic dimensions, such as articulation and phonation. Relevant breakthroughs could be made, under standardized recording conditions, by comparing the predominance of different motor speech alterations in each nfvPPA subgroup, as done elsewhere.^8, 45^ Third, our approach could be more stringently tested through comparisons between nfvPPA and lvPPA, whose clinical differentiation proves particularly challenging when using standard speech measures.^13,15^ Also, post-mortem confirmation for svPPA participants would be useful to refine pathology-related conclusions. Fourth, while the MMSE was statistically preferable to other severity measures for covariance analyses, it relies heavily on speech and language functions. In this sense, each group mainly missed different items (sentence comprehension, repetition, and writing for nfvPPA-CBD and nfvPPA-PSP; object naming and backward spelling for svPPA). Future studies could further explore how speech timing is affected by different language profiles and employ relevant, non-verbal measures for covariance analyses. Fifth, since data was collected through various protocols over 17 years, we lacked a single, objective grammatical measure across patients. While this precludes detailed characterizations of the patients’ aphasic profile, future research could use morphosyntactic analyses of other recorded samples to elucidate the issue. Sixth, our cohort included too few persons with Pick’s disease, preventing their inclusion for statistical analysis and inviting new research on their particular speech timing profiles. Finally, our approach could be implemented on platforms offering remote audio recording. Free open-source applications already provide user-friendly capabilities for clinical research. These tools could foster continual remote testing, opening exciting opportunities for longitudinal research and disease progression monitoring.

## Conclusion

Automated speech timing measures can discriminate persons with autopsy-confirmed nfvPPA from HCs and patients with svPPA, identify persons with autopsy-confirmed nfvPPA at the probabilistic single-person level, capture distinct atrophy patterns across syndrome-sensitive regions, and potentially differentiate between patients with CBD and PSP pathology. Computerized speech tools could, thus, contribute to differential diagnosis and pathology prediction in this population. New avenues can be envisaged towards developing scalable, objective innovations in clinicopathological FTLD assessments.

## Supporting information

eAppendix

## Conflicts of interest

None to declare.

## Funding

A. M. García is an Atlantic Fellow at the Global Brain Health Institute (GBHI) and is supported with funding from GBHI, Alzheimer’s Association, and Alzheimer’s Society (GBHI ALZ UK-22-865742); as well as CONICET; ANID (FONDECYT Regular 1210176); and Programa Interdisciplinario de Investigación Experimental en Comunicación y Cognición (PIIECC), Facultad de Humanidades, USACH. A. E. Welch reports no disclosures relevant to the manuscript. M. L. Mandelli reports no disclosures relevant to the manuscript. M. L. Henry is supported by the National Institutes of Health (NIDCD R01DC016291). S. Lukic reports no disclosures relevant to the manuscript. M. J. Torres-Prioris has been funded by a postdoctoral fellowship under the program Plan Andaluz de Investigación, Desarrollo e Innovación (PAIDI 2020) (DOC_00421). J. Deleon is supported by the National Institutes of Health (NIDCD K23 DC018021). B. M. Ratnasiri reports no disclosures relevant to the manuscript. D. L. Lorca-Puls is supported by a postdoctoral fellowship from the National Agency for Research and Development (ANID BECAS-CHILE 74200073). B. L. Miller reports no disclosures relevant to the manuscript. W. W. Seeley reports no disclosures relevant to the manuscript. A. P. Vogel is Chief Science Officer of Redenlab Inc. The Memory and Aging Center is also supported by the Larry L. Hillblom Foundation; John Douglas French Alzheimer’s Foundation; Koret Family Foundation; Consortium for Frontotemporal Dementia Research; and McBean Family Foundation. Maria Luisa Gorno-Tempini is supported by grants from the National Institutes of Health (NINDS R01 NS050915, NIDCD K24 DC015544; NIA U01 AG052943).

## Acknowledgments

We thank all participants and their families for contributing to this research.

## Notes

### Competing Interest Statement

The authors have declared no competing interest.

### Funding Statement

A. M. Garcia is an Atlantic Fellow at the Global Brain Health Institute (GBHI) and is supported with funding from GBHI, Alzheimer's Association, and Alzheimer's Society (GBHI ALZ UK-22-865742); as well as CONICET; ANID (FONDECYT Regular 1210176); and Programa Interdisciplinario de Investigacion Experimental en Comunicacion y Cognicion (PIIECC), Facultad de Humanidades, USACH. A. E. Welch reports no disclosures relevant to the manuscript. M. L. Mandelli reports no disclosures relevant to the manuscript. M. L. Henry is supported by the National Institutes of Health (NIDCD R01DC016291). S. Lukic reports no disclosures relevant to the manuscript. M. J. Torres-Prioris has been funded by a postdoctoral fellowship under the program Plan Andaluz de Investigacion, Desarrollo e Innovacion (PAIDI 2020) (DOC_00421). J. Deleon is supported by the National Institutes of Health (NIDCD K23 DC018021). B. M. Ratnasiri reports no disclosures relevant to the manuscript. D. L. Lorca-Puls is supported by a postdoctoral fellowship from the National Agency for Research and Development (ANID BECAS-CHILE 74200073). B. L. Miller reports no disclosures relevant to the manuscript. W. W. Seeley reports no disclosures relevant to the manuscript. A. P. Vogel is Chief Science Officer of Redenlab Inc. The Memory and Aging Center is also supported by the Larry L. Hillblom Foundation; John Douglas French Alzheimer's Foundation; Koret Family Foundation; Consortium for Frontotemporal Dementia Research; and McBean Family Foundation. Maria Luisa Gorno Tempini is supported by grants from the National Institutes of Health (NINDS R01 NS050915, NIDCD K24 DC015544; NIA U01 AG052943).

