## Supplementary material for "Automated detection of speech timing alterations in autopsy-confirmed non-fluent/agrammatic variant primary progressive aphasia": eAppendix

**Supplement**

**eAppendix 1. Neurological characterization of each participant group**

Following enrollment at USCF’s Memory and Aging Center, all patients were phenotypically diagnosed with a PPA variant based on multidisciplinary evaluations considering clinical presentation, validated tests, and experimental assessment results. At first presentation, beyond their primary speech and language deficits, patients with nfvPPA tended to present with executive dysfunction and spared visuospatial skills. Neurological examination revealed parkinsonian features in most patients, including asymmetric (R > L) tremor, bradykinesia, and oculomotor initiation or saccadic difficulties. There were no neurological findings that consistently distinguished persons with nfvPPA-CBD from those with nfvPPA-PSP. Both groups were characterized by oculomotor and gait abnormalities, behavioral impulsivity, and rigidity. Tremor, non-speech apraxia, and agraphesthesia were more common in nfvPPA-CBD patients, whereas swallowing, ocular pursuit, and saccadic difficulties were more frequent in nfvPPA-PSP patients. No signs of balint syndrome or simultagnosia were reported in either group. In patients with svPPA, cognitive tests revealed mental rigidity and occasional socio-semantic behavioral changes with spared episodic and working memory. Neurologically, they presented with substantially less motor and parkinsonian findings. Finally, HCs had MMSE scores of 28-30 and no motor, cognitive or linguistic findings suggestive of neurodegenerative disease.

**eAppendix 2. Clinical information on the nfvPPA-CBD and nfvPPA-PSP subgroups**

While some patients with nfvPPA may have eventually presented with corticobasal syndrome or progressive supranuclear palsy syndrome, there were no clear differential signs of either syndrome in either subgroup at the time of testing (eTable 1). In line with reported criteria,^e1^ some patients with nfvPPA-CBD exhibited, during the course of their disease, (1) slowly progressive course; (2) asymmetric limb or axial rigidity, present without reinforcement; (3) aphasia, visuospatial impairment or neglect, or apraxia; and (4) dystonia, myoclonus, cortical sensory loss, or alien limb phenomenon. Some patients in the nfvPPA-PSP group presented with or developed (1) a gradually progressive disorder with onset at age 40 or later; and (2) vertical supranuclear gaze palsy and prominent postural instability. Crucially, however, predominant signs of each subgroup were partly observed in the other. Also, in previous work encompassing many of this study’s patients,^e1^ nfvPPA-CBD patients exhibited a trend toward greater sentence comprehension deficits, whereas nfvPPA-PSP participants presented greater signs of dysarthria and depression. Moreover, neurological reports offered no unambiguous ground for predicting pathology in either nfvPPA group, partly because data was collected over approximately 17 years, during which time there were substantial changes in neurological protocols and overall understanding of clinico-pathological correlations.

Longitudinally, both groups of patients developed unintelligible and agrammatic speech, with spared naming and semantic abilities as well as parkinsonian limb and eye movement abnormalities. The latest neurological reports for patients in the nfvPPA-CBD group more frequently mentioned alien limb phenomenon and limb apraxia as later occurring symptoms, while those from the nfvPPA-PSP cohort mentioned falls and dysphagia. For a description of the neurological and cognitive progression of a similar cohort of autopsy-confirmed patients with nfvPPA-CBD and nfvPPA-PSP, see Santos-Santos et al. (2016).^e^^1^

**eTable 1.** Neurological features of the nfvPPA subgroups.

|  | **Patients with nfvPPA-CBD**  **(*n* = 12)** | **Patients with nfvPPA-PSP**  **(*n* = 10)** | ***p*-value** |
| --- | --- | --- | --- |
| **Neurological symptoms** | | | |
| Ocular movements  (0-4; 4 = most impaired) | 0.4 (0.6) | 0.6 (0.5) | 0.5 |
| Myoclonus (% present)  (0 = normal; 1 = abnormal) | 0 (0) | 0 (0) | ----- |
| Motor tone  (0-4; 4 = most impaired) | 0.7 (0.5) | 0.8 (0.4) | 0.8 |
| Dystonia  (0-4; 4 = most impaired) | 0.3 (0.5) | 0.1 (0.3) | 0.4 |
| Tremor  (0-4; 4 = most impaired) | 0.6 (0.7) | 0.3 (0.5) | 0.3 |
| Non-speech praxia  (0-2; 2 = most impaired) | 0.7 (0.9) | 0.6 (0.7) | 0.7 |
| Cortical sensory function (% present)  (0 = normal; 1 = abnormal) | 63.6 | 66.7 | .28 |
| Gait (% present)  (0 = normal; 1 = abnormal) | 81% | 100% | .003 |
| Comparisons of categorical variables (myoclonus, cortical sensory function, and gait scores) were performed via χ2 tests. Every other variable was compared with two-tailed, unpaired *t*-tests. NfvPPA-CBD: non-fluent variant primary progressive aphasia with corticobasal degeneration pathology; NfvPPA-PSP: non-fluent variant primary progressive aphasia with progressive supranuclear palsy pathology. | | | |

**eAppendix 3. Participant information for the four-group setting**

**eTable 2.** Participants’ demographic, speech, language, and cognitive profiles.

|  | **Patients with nfvPPA-CBD**  **(*n* = 12)** | **Patients with nfvPPA-PSP**  **(*n* = 10)** | **Patients with svPPA**  **(*n* = 15)** | **Healthy controls**  **(*n* = 10)** | ***p*-value** |
| --- | --- | --- | --- | --- | --- |
| **Demographics** | | | | | |
| % Male | 33.3% | 50% | 46.7% | 50% | .831 |
| % Right-handed | 100% | 90% | 93.3% | 80% | .406 |
| Age | 64.6 (7.3) | 70.6 (5.6) | 66.5 (5.8) | 70 (6.9) | .057 |
| Years of education | 15.9 (3.1) | 17.3 (3.3) | 17.9 (2.4) | 17 (1) | .434 |
| **Disease duration** |  |  |  |  |  |
| Years post onset | 3.3 (0.9) | 4.1 (1.5) | 5.6 (3.3) | ----- | .112 |
| **Perceptual speech assessment** | | | | | |
| Apraxia of speech severity  (1 = minimal, 7 = profound) | 1.6 (1.1)^c,d^ | 1.6 (1.8)^c,d^ | 0 (0)^a,b^ | 0 (0)^a,b^ | < .001* |
| Dysarthria severity  (1 = minimal, 7 = profound) | 1.5 (1.5)^c,d^ | 2.3 (1.6)^c,d^ | 0 (0)^a,b^ | 0 (0)^a,b^ | < .001* |
| **Language measures** | | | | | |
| Boston Naming Test (15 items) | 12.5 (3.0)^c^ | 13 (1.6)^c^ | 6.2 (3.8)^a,b,d^ | 14.7 (0.5)^c^ | < .001* |
| WAB spontaneous speech fluency (max score: 10) | 6.9 (2.1)^c^ | 7.6 (1.7) | 9.1 (0.6)^a^ | ----- | .012* |
| Syntactic processing (% correct from Wilson et al., 2010)^e^^2^ | 88.4 (14.4) | 97 (3.3) | 96.1 (4.8) | 99.4 (1.5) | .141 |
| **Cognitive measures** | | | | | |
| CDR (sum of boxes) | 0.6 (0.2)^d^ | 0.4 (0.3)^d^ | 0.6 (0.2)^d^ | 0 (0)^a,b,c^ | < .001* |
| Language | 1.54 (0.5)^c^ | 1.33 (0.7) | 1 (0.3)^a^ | ----- | .036* |
| FTLD CDR | 5 (1.9)^d^ | 3.5 (2)^d^ | 5 (2.1)^d^ | 0 (0)^a,b,c^ | < .001* |
| MMSE | 24.5 (5.2)^d^ | 26.2 (3.4) | 25.7 (3.4) | 29.4 (0.8)^a^ | < .001* |
| Digits forwards | 4.5 (0.8)^c^ | 4.8 (1.3) | 6.3 (1.4)^a^ | ----- | .017* |
| Digits backwards | 2.9 (0.6)^c^ | 4.0 (1.3) | 4.7 (1.2)^a^ | ----- | .002* |
| Design fluency | 6.5 (3.3) | 6.8 (3.1) | 9.6 (2.8) | ----- | .020* |
| Benson recall | 10.5 (3.7) | 9.7 (4.0) | 9.8 (3.3) | ----- | .856 |
| GDS | 7.3 (4.6) | 7.1 (8.5) | 7.8 (6.0) | ----- | .662 |
| Asterisks (*) indicate significant differences at *p* < .05. Demographic information and performance between groups were compared via ANOVAs. Superscript letters indicate the group showing significant differences in the post-hoc pairwise comparisons (via Tukey’s HSD tests). a: nfvPPA; b: svPPA; c: healthy controls. Sex and handedness were compared with χ2 tests. NfvPPA-CBD: non-fluent variant primary progressive aphasia with corticobasal degeneration pathology; NfvPPA-PSP: non-fluent variant primary progressive aphasia with progressive supranuclear palsy pathology; svPPA: semantic variant primary progressive aphasia; WAB: Western Aphasia Battery; FTLD CDR: Frontotemporal Lobar Degeneration Clinical Dementia Rating; MMSE: Mini-Mental State Examination; GDS: Geriatric Depression Scale. | | | | | |

**eAppendix 4. Power estimation**

We ran power estimation analyses on G*Power 3.1^e^^3^ to establish the minimal sample size required for the three-group (nfvPPA, HCs, svPPA) and the four-group analyses (nfvPPA-CBD, nfvPPA-PSP, HCs, svPPA). In both cases, we considered one-way ANOVAs, alpha of .05, an effect size of η_p_^2^ = 0.25, and a power of 0.8. Results indicated that robust effects could be detected with 33 participants for the three-group approach and 40 for the four-group approach. Our actual sample size (*n* = 47) reaches a power of 0.93 for the three-group setting and 0.90 for the four-group setting.

**eAppendix 5. Text used for the overt reading task^e4^**

**Grandfather Passage:** You wish to know all about my grandfather. Well, he is nearly 93 years old, yet he still thinks as swiftly as ever. He dresses himself in an old black frock coat, usually several buttons missing. A long beard clings to his chin, giving those who observe him a pronounced feeling of the utmost respect. When he speaks, his voice is just a bit cracked and quivers a bit. Twice each day he plays skillfully and with zest upon a small organ. Except in the winter when the snow or ice prevents, he slowly takes a short walk in the open air each day. We have often urged him to walk more and smoke less, but he always answers, “Banana oil!” Grandfather likes to be modern in his language.

**eAppendix 6. Supplementary ANCOVA results**

**eTable 3.** Results for the three-group setting (all patients with nfvPPA together).

|  | **Patients with nfvPPA** | **Patients with svPPA** | **Healthy controls** | **Main effect** | | | |
| --- | --- | --- | --- | --- | --- | --- | --- |
|  | **Mean (*SD*)** | | |  | ***F*** | ***p*-value** | $\boldsymbol{\eta}_{\mathbf{p}}^{\mathbf{2}}$ |
| **Articulation rate**  (syllables per phonated second) | 2 (0.6)^b,c^ | 2.9 (0.5)^a^ | 3.2 (0.4)^a^ |  | 16.654 | **< .001*** | 0.442 |
| **Mean syllable duration**  (seconds) | 0.5 (0.1)^b,c^ | 0.4 (0.1)^a^ | 0.3 (0.04)^a^ |  | 14.454 | **< .001*** | 0.408 |
| **Mean pause duration**  (seconds) | 0.4 (0.2)^b,c^ | 0.2 (0.1)^a^ | 0.2 (0.02)^a^ |  | 9.473 | **< .001*** | 0.316 |
| **Syllable duration variability**  (seconds) | 0.5 (0.2)^b,c^ | 0.2 (0.1)^a^ | 0.2 (0.04)^a^ |  | 8.236 | **< .001*** | 0.287 |
| **Pause duration variability**  (seconds) | 0.4 (0.3)^b,c^ | 0.2 (0.1)^a^ | 0.2 (0.04)^a^ |  | 6.828 | **.003*** | 0.250 |
| Superscript letters indicate the group showing significant differences in post-hoc pairwise comparisons, via Tukey’s HSD tests. a: nfvPPA; b: svPPA; c: Healthy controls. Asterisks (*) indicate significant differences at *p* < .05. NfvPPA: non-fluent variant primary progressive aphasia; svPPA: semantic variant primary progressive aphasia. | | | | | | | |

**eTable 4.** Results for the four-group setting.

|  | **Patients with nfvPPA-CBD** | **Patients with nfvPPA-PSP** | **Patients with svPPA** | **Healthy controls** | **Main effect** | | |
| --- | --- | --- | --- | --- | --- | --- | --- |
| **Variable** | **Mean (*SD*)** | | | | ***F*** | ***p*-value** | $\boldsymbol{\eta}_{\mathbf{p}}^{\mathbf{2}}$ |
| **Articulation rate** (syllables per phonated second) | 1.7 (0.3)^b,c,d^ | 2.4 (0.6)^a,d^ | 2.9 (0.5)^a^ | 3.2 (0.4)^a,b^ | 16.88 | **< .001*** | 0.553 |
| **Mean syllable duration**  (seconds) | 0.6 (0.1)^b,c,d^ | 0.4 (0.2)^a^ | 0.4 (0.1)^a^ | 0.3 (0.04)^a^ | 15.67 | **< .001*** | 0.534 |
| **Mean pause duration**  (seconds) | 0.4 (0.1)^b,c,d^ | 0.2 (0.1)^a^ | 0.2 (0.1)^a^ | 0.2 (0.02)^a^ | 19.00 | **< .001*** | 0.594 |
| **Syllable duration variability**  (seconds) | 0.6 (0.2)^b,c,d^ | 0.3 (0.2)^a^ | 0.2 (0.1)^a^ | 0.2 (0.04)^a^ | 10.63 | **< .001*** | 0.444 |
| **Pause duration variability**  (seconds) | 0.6 (0.3)^b,c,d^ | 0.3 (0.1)^a^ | 0.2 (0.1)^a^ | 0.2 (0.04)^a^ | 9.01 | **< .001*** | 0.403 |
| Superscript letters indicate the group showing significant differences in the post-hoc pairwise comparison, via Tukey’s HSD tests. a: nfvPPA-CBD; b: nfvPPA-PSP; c: svPPA; d: healthy controls. Asterisks (*) indicate significant differences at *p* < .05. NfvPPA-CBD: non-fluent variant primary progressive aphasia with corticobasal degeneration pathology; nfvPPA-PSP: non-fluent variant primary progressive aphasia with progressive supranuclear palsy pathology; svPPA: semantic variant primary progressive aphasia. | | | | | | | |

**eAppendix 7. Correlations between automated and perceptual motor speech measures**

**eTable 5.** Pearson’s correlations between automated and perceptual motor speech measures in the nfvPPA group.

|  | **Articulation rate**  (syllables per phonated second) | **Mean syllable duration**  (seconds) | **Mean pause duration**  (seconds) | **Syllable duration variability**  (seconds) | **Pause duration variability**  (seconds) |
| --- | --- | --- | --- | --- | --- |
| **Apraxia**  **of speech** | *p* = .04  *r* = -0.45 | *p* = .04  *r* = 0.46 | *p* = .19  *r* = .31 | *p* = .32  *r* = 0.23 | *p* = .53  *r* = 0.14 |
| **Dysarthria** | *p* = .49  *r* = -0.16 | *p* = .83  *r* = 0.05 | *p* = .90  *r* = 0.03 | *p* = .76  *r* = 0.07 | *p* = .83  *r* = 0.05 |

e.4. Van Riper C. *Speech Correction: Principles and Methods*, fourth edition. Englewood Cliffs, NJ: Prentice-Hall. 1963.
